# A splicing-based multi-tissue joint transcriptome-wide association study identifies susceptibility genes for breast cancer

**DOI:** 10.1101/2023.10.15.23297045

**Authors:** Guimin Gao, Julian McClellan, Alvaro N. Barbeira, Peter N. Fiorica, James L. Li, Zepeng Mu, Olufunmilayo I. Olopade, Dezheng Huo, Hae Kyung Im

**Affiliations:** Department of Public Health Sciences, University of Chicago, IL, 60637, USA; Section of Genetic Medicine, Department of Medicine, University of Chicago, IL, 60637, USA; Section of Hematology & Oncology, Department of Medicine, University of Chicago, IL, 60637, USA

**Keywords:** alternative splicing, transcriptome-wide association studies, breast cancer, intron excision, multi-tissue, joint analysis, susceptibility genes

## Abstract

Splicing-based transcriptome-wide association studies (splicing-TWASs) of breast cancer have the potential to identify new susceptibility genes. However, existing splicing-TWASs test association of individual excised introns in breast tissue only and have thus limited power to detect susceptibility genes. In this study, we performed a multi-tissue joint splicing-TWAS that integrated splicing-TWAS signals of multiple excised introns in each gene across 11 tissues that are potentially relevant to breast cancer risk. We utilized summary statistics from a meta-analysis that combined genome-wide association study (GWAS) results of 424,650 European ancestry women. Splicing level prediction models were trained in GTEx (v8) data. We identified 240 genes by the multi-tissue joint splicing-TWAS at the Bonferroni corrected significance level; in the tissue-specific splicing-TWAS that combined TWAS signals of excised introns in genes in breast tissue only, we identified 9 additional significant genes. Of these 249 genes, 88 genes in 62 loci have not been reported by previous TWASs and 17 genes in 7 loci are at least 1 Mb away from published GWAS index variants. By comparing the results of our spicing-TWASs with previous gene expression-based TWASs that used the same summary statistics and expression prediction models trained in the same reference panel, we found that 110 genes in 70 loci identified by our splicing-TWASs were not reported in the expression-based TWASs. Our results showed that for many genes, expression quantitative trait loci (eQTL) did not show significant impact on breast cancer risk, while splicing quantitative trait loci (sQTL) showed strong impact through intron excision events.

## Introduction

Breast cancer is a complex genetic disorder caused by high-penetrance genes, multiple common variants, and non-genetic factors (i.e. environmental and lifestyle/reproductive factors). To date, genome-wide association studies (GWASs) have identified over 200 loci significantly associated with breast cancer. However, the common susceptibility variants identified by GWASs account for a relatively small proportion of the familial relative risk (1) and specific causal genes in most of these loci have not been identified. To further explore the role of genetic variants on breast cancer, expression-based transcriptome-wide association studies (expression-TWASs) have identified hundreds of genes whose genetically regulated gene expression is significantly associated with breast cancer and its various subtypes (2–5). While expression-TWAS have contributed to our understanding of genetic risk in breast cancer, research has recently shown that RNA splicing is major contributor to complex traits. In fact, splicing quantitative trait loci (sQTLs) and expression quantitative loci (eQTLs) may both have significant effects on phenotypes (6). Splicing-based TWASs (splicing-TWASs) can use splicing information of individual intron excision events (i.e., the read proportions for individual introns within an alternatively excised intron cluster in RNA-seq data), which often cannot be captured by traditional expression-based TWASs that use the total gene expression levels in a gene. Thus, investigating alternative splicing in the context of breast cancer may help identify new susceptibility genes. Indeed, recent studies suggest that alternative splicing plays a critical role in the genetic regulation of breast cancer (7) and a splicing-TWAS has identified 85 genes associated with breast cancer by applying a susceptible transcription factor (sTF)-TWAS method to alternative splicing levels measured at individual excised introns in breast tissue (8). However, existing splicing-TWASs suffer from limited power from the multiple testing correction required to account for the large number of tests used to determine the association individual excised introns with disease phenotypes. We hypothesize the power of splicing-TWAS could be increased by modeling multiple intron excision splicing events jointly—a reasonable approach since the multiple (intron excision) splicing events in a gene may jointly impact the phenotype. Existing splicing-TWASs for breast cancer are also limited by their use breast tissue only. Other tissues potentially relevant to breast cancer development may provide useful information in the identification of genes susceptible to breast cancer (9) and including multiple tissues in splicing-TWASs could prove fruitful.

In this study, we performed a joint splicing-TWAS that combine information from multiple excised introns in each gene across multiple tissues that are potentially relevant to breast cancer. We used intron splicing level prediction models for 11 tissues relevant to breast cancer trained in GTEx v8 data (10, 11). We used summary statistics from a meta-analysis of the GWAS results from the Breast Cancer Association Consortium (BCAC) (including 122,977 breast cancer cases and 105,974 controls) (1) and GWAS results of 10,534 breast cancer cases and 185,116 controls in UK Biobank (UKB) (12). Finally, we compared results from our current joint splicing-TWAS with those of the previous joint expression-TWAS, which uses the same GWAS summary statistics as the splicing-TWAS and for which the gene expression prediction models are trained in the same 11 tissues in GTEx v8 data.

## Results

### Joint splicing-TWAS combines information from multiple intron excision events in a gene across multiple tissues

To create our multi-tissue joint splicing-TWAS, we used summary statistics from a meta-analysis (5) that combined GWAS results on women of European ancestry from the Breast Cancer Association Consortium (BCAC) (1) and GWAS results on European women from UK Biobank (UKB) (5). We used splicing level prediction models for 11 selected tissues potentially relevant to breast cancer (5) (see also Methods). The models were trained on samples with European ancestry from GTEx v8 data (sample sizes ranging from 129 to 670 with a median of 227) using a multivariate adaptive shrinkage (MASH) method (10, 13, 14). In total, we tested 14,528 genes across the 11 tissues with splicing prediction models, including 10,931 genes in breast tissue in our splicing TWAS analysis. In the prediction models, splicing levels were quantified at clustered excised introns using short-read RNA-seq data by LeafCutter (15). Specifically, read proportions for the introns in each cluster were estimated and quantile normalized.

In our joint splicing-based TWAS analysis, we analyzed genes with splicing prediction models for at least one intron (excision) event. First, we performed a traditional individual intron-based TWAS analysis for all intron excision events in all genes across the genome in each tissue by the S-PrediXcan method (16). Second, for each gene with prediction models for multiple intron excision events, we combined the S-PrediXcan p-values for the multiple introns in each tissue by the aggregated Cauchy association test (ACAT) method (17) to obtain a tissue-specific gene-level p-value. Finally, for the multi-tissue TWAS, we combined the tissue-specific p-values across the 11 selected tissues to generate an overall p-value for each gene using ACAT; we ignored tissues that had no prediction models and, therefore, no p-values.

Of the 14,528 genes tested in our multi-tissue joint splicing-TWAS analysis, we identified 240 genes (located in 94 loci) at the Bonferroni corrected significance level (p < 3.44 X 10^-6^) to be associated with breast cancer (Supplementary Table 1). We also created a breast-tissue–specific joint splicing-TWAS using the ACAT method to combine multiple excised introns in a gene in breast tissue only, we identified 158 genes significant at the Bonferroni corrected significance level (p < 4.57 X 10^-6^), of which 149 genes were also identified by the multi-tissue joint splicing-TWAS that combined information across 11 tissues (Supplementary Table 2). A total of 249 significant genes that were identified by either the multi-tissue or breast-tissue–specific joint splicing-TWAS. Of these 249 genes, 88 have not been reported by previous TWASs (Supplementary Table 1). We found 17 genes in 7 loci that are at least 1 Mb away from previously published GWAS index variants, including 11 genes in 7 loci not reported by previous TWASs (Table 1). Of the 17 genes in Table 1, 12 were identified by both the multi-tissue and breast-tissue-specific joint splicing TWASs and two genes (*AFF1* and *SRP54*) were identified only by the breast-tissue-specific joint splicing-TWAS. We further performed conditional splicing-TWAS tests for the significant genes adjusting for nearby GWAS index risk variants (within ±2 Mb of the transcription start or stop sites of a gene). The 17 genes remained significant in the conditional splicing-TWASs (see “Conditional ACAT P-value” in Supplementary Table 1 and more details in Methods); this suggests the TWAS signals at the 17 genes are independent of nearby GWAS index variants. Our results also suggest that the multi-tissue, joint splicing-TWAS, especially when used in tandem with breast-tissue–specific joint splicing-TWAS, could provide additional information regarding disease susceptibility genes than GWAS or expression-TWAS could provide alone.

**Table 1.** The 17 genes identified by splicing-TWASs located at 7 loci at least 1 Mb away from previous GWAS hits.

| Cytoband | Gene symbol | Gene position (hg38) | Gene type (v40) | Multi-Tissue ACAT P<sup>1</sup> | Breast-tissue ACAT P<sup>2</sup> | Max PIP<sup>3</sup> | MAX RCP<sup>4</sup> | Reported in previous TWASs |
| --- | --- | --- | --- | --- | --- | --- | --- | --- |
| 1q21.1 | FCGR1CP | 1:143874793-143883575 | pseudogene | 1.90E-09 | NA | 0.994 | 0 | No |
| 4q21.3 | **AFF1\*** | 4:86935002-87141039 | protein_coding | 1.33E-05 | **3.58E-06** | 0.659 | 0.001 | No |
| 6q24.1 | TXLNB | 6:139240061-139291998 | protein_coding | 1.87E-06 | NA | 0.56 | 0.364 | Yes |
|  | ENSG00000226571 | 6:139271362-139667284 | lncRNA | 2.24E-06 | NA | 0.76 | 0.452 | No |
| 7q22.1 | TRIM4 | 7:99876958-99919531 | protein_coding | 6.51E-07 | 4.55E-07 | 0.0917 | 0.065 | No |
|  | GJC3 | 7:99923266-99929620 | protein_coding | 1.06E-06 | 8.23E-07 | 0.0577 | 0.002 | No |
|  | AZGP1 | 7:99966720-99976042 | protein_coding | 1.06E-06 | 8.23E-07 | 0.0577 | 0.002 | No |
|  | PMS2P1 | 7:100300024-100336307 | pseudogene | 1.90E-07 | 2.02E-07 | 0.0604 | 0.077 | No |
|  | STAG3L5P | 7:100336079-100351900 | pseudogene | 2.52E-07 | 7.94E-07 | 0.08 | 0.235 | No |
|  | PILRB | 7:100352176-100367831 | protein_coding | 2.38E-07 | 2.36E-07 | **0.144** | 0.258 | Yes |
|  | PILRA | 7:100367530-100400096 | protein_coding | 1.15E-06 | 7.50E-07 | 0.0277 | 0.087 | Yes |
|  | ZCWPW1 | 7:100400826-100428992 | protein_coding | 3.43E-07 | 3.57E-07 | 0.0566 | 0.104 | Yes |
|  | TSC22D4 | 7:100463359-100479232 | protein_coding | 4.23E-07 | 3.42E-07 | 0.0412 | 0.075 | Yes |
|  | NYAP1 | 7:100483927-100494802 | protein_coding | 5.73E-07 | NA | 0.017 | 0.016 | Yes |
| 14q13.2 | **SRP54\*** | 14:34981957-35029686 | protein_coding | 6.71E-06 | **1.63E-06** | 0.684 | 0.379 | No |
|  | PRORP | 14:35121846-35277622 | protein_coding | 3.53E-06 | 3.75E-06 | 0.774 | 0.134 | No |
| 17p12 | ZNF18 | 17:11977439-11997475 | protein_coding | 2.98E-06 | 1.24E-06 | 0.905 | 0.456 | No |

\* These two genes in bold are significant in the ACAT test in breast tissue but marginally significant in ACAT test across 11 tissues.
<sup>1</sup> P value of multi-tissue ACAT test that combined multiple intron-based TWAS p-values in a gene across 11 tissues.
<sup>2</sup> P value of ACAT test that combined multiple intron-based TWAS p-values in a gene in breast tissue only.
<sup>3</sup> Maximum PIP for all introns across 11 tissues.
<sup>4</sup> Maximum RCP for all introns across 11 tissues.
Abbreviation: GWAS, genome-wide association study; TWAS, transcriptome-wide association study; ACAT, aggregated Cauchy association test; PIP, posterior inclusion probability; RCP, regional colocalization probability.

Of the 17 genes in Table 1 that were identified by our current splicing-TWASs and are at least 1 Mb away from previous GWAS hits, 11 genes have not been identified by any previous published TWASs (Table 2). For each of these genes, we further explored which tissues and which specific excised introns in the tissues had the strongest signals. Table 2 lists the tissue-specific joint splicing-TWAS p-values for breast tissue and/or the tissues with strongest tissue-specific signals; in addition, Table 2 lists the S-PrediXcan z-scores and p-values for excised introns in these tissues. For example, at the gene *FCGR1CP*, there were four intron events with prediction models in whole blood tissue, and the *FCGR1CP* isoforms with introns excised from 143,874,823 to 143,875,219 base pairs in chromosome 1 (intron_1_143874823_143875219) had the smallest p-value (4.75 X 10^-10^); however, in breast tissue, no excised intron events had prediction models (i.e., intron phenotypes could not be predicted by sQTL variants). Furthermore, In Table 2, 9 of the 11 genes showed significant associations with breast cancer in breast tissue (i. e. with tissue specific splicing-TWAS p-values < 4.57×10^-6^), indicating these genes potentially impact breast cancer risk by splicing in breast tissue.

**Table 2.**
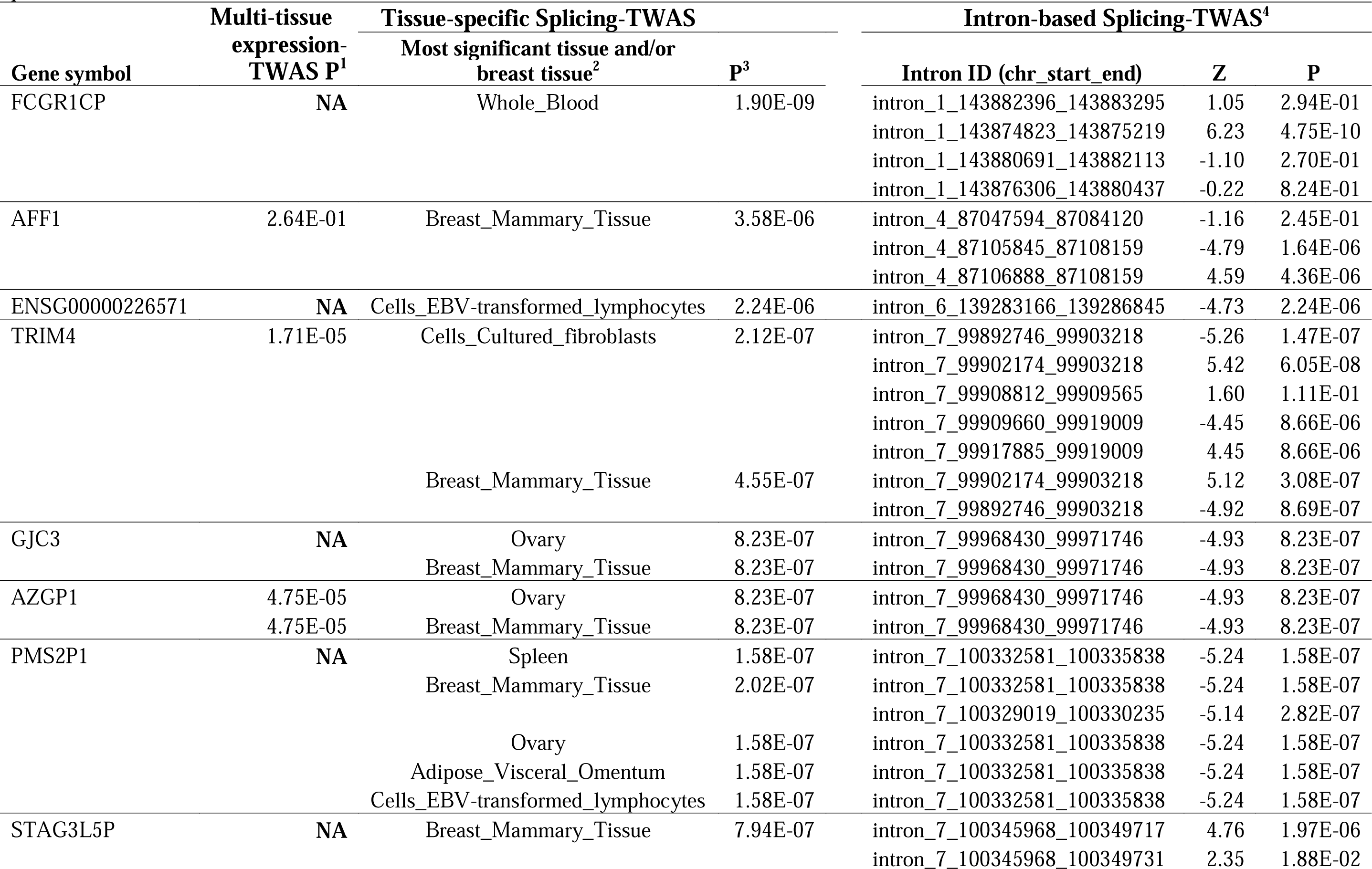
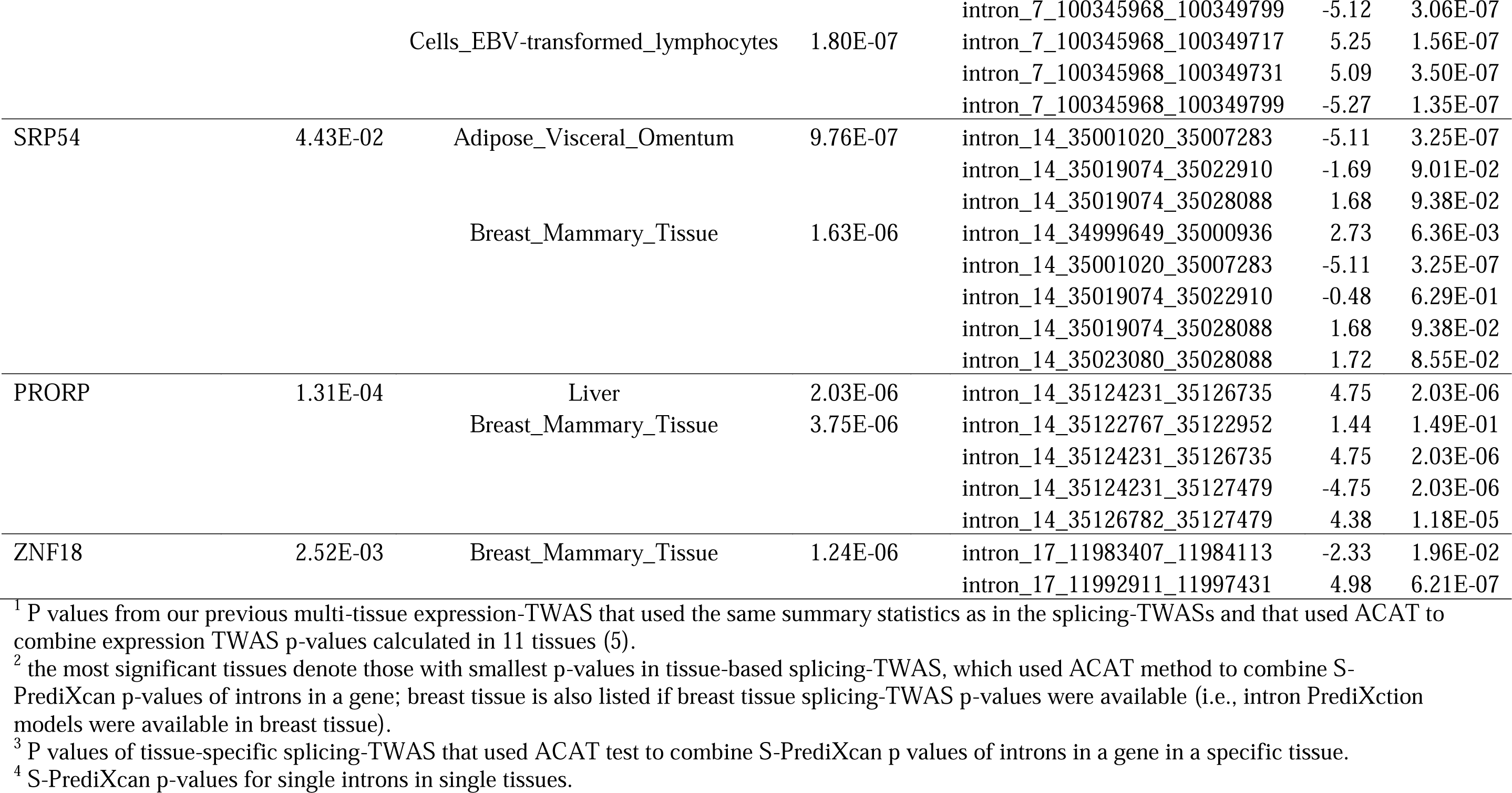
The 11 genes identified by splicing-TWASs and were at least 1 Mb away from previous GWAS hits but not identified by previous TWASs.

### Joint splicing-TWASs identify 110 genes not found by expression-TWASs that used the same GWAS summary statistics

We previously performed both a multi-tissue and a breast-tissue-specific expression-TWASs that used the same meta-analysis summary statistics as described above and expression prediction models trained in the same 11 tissues (or breast tissue only) from GTEx v8 (5). We compared the combined results of our current multi-tissue and breast-tissue–specific joint splicing-TWASs with the combined results from our previous multi-tissue and breast-tissue–specific expression-TWASs. The expression-TWASs identified 309 genes (5)compared to 249 identified by splicing-TWASs. The splicing-TWASs and expression-TWASs mutually identified 139 genes; the remaining 110 and 170 genes were unique to splicing-TWASs and expression-TWASs, respectively. Supplementary Table 3 lists the comparison of p-values and/or z-scores of the two types of (expression- and splicing-based) TWASs for the 110 genes unique to joint splicing-TWASs. For 83 genes, the expression-TWASs had weak signals but did not reached the Bonferroni corrected significance level (p ≤ 2.59×10^-6^). For the remaining 27 genes, no expression prediction models were available in the 11 tissues considered (Supplementary Table 3, genes with “NA” in the column “Expression-TWAS ACAT joint P-value”); because eQTL signals in the reference panel (GTEx V8) were insufficient in cis-gene regions to predict the gene expression levels. While the eQTLs in these cis-gene regions did not show significant impact on breast cancer risk through gene expression in our data analyses, prediction models for excised introns of these genes were created in at least one of the 11 tissues in the reference panel and the splicing-TWASs showed significance in these genes. This evidence suggests sQTL in these cis regions have strong impact on breast cancer risk through the excised introns. As a special case, in Table 2 we also included p-values of our previous multi-tissue expression-TWAS for the 12 genes that were not identified by our expression-TWASs but identified by our splicing-TWASs and are at least 1Mb away from GWAS index SNPs. For five genes (*FCGR1CP*, *ENSG00000226571*, *GJC3*, *PMS2P1*, *STAG3L5P*), we found no available expression prediction models in the 11 tissues but having intron splicing prediction models in at least one of the 11 tissues.

### Intron-based fine mapping identifies 114 genes with high posterior inclusion probability

Our splicing-TWASs were based on S-PrediXcan (16) that were applied to test intron–trait association for individual excised introns in single tissues. The intron–trait association statistics for introns in a linkage disequilibrium (LD) region can be correlated as a function of LD among genetic variants and sQTL weights. As a result, when a causal intron is associated with breast cancer, S-PrediXcan may identify significant intron-trait associations at a set of introns in the LD region, including non-causal introns. To narrow down the list of potential causal genes from the 249 genes identified by our splicing-TWASs, we performed intron-based fine mapping on a set of excised introns in an LD region in each tissue using the package FOCUS (18). We calculated marginal posterior inclusion probability (PIP) for each intron. For a gene, we calculated maximum PIP (Max-PIP) for introns in the gene across 11 tissues. We considered genes with a Max-PIP > 0.80 to have a high likelihood of being causal. In our fine-mapping analysis, 114 genes had Max-PIP greater than 0.80 (Supplementary Table 4).

### Colocalization combined with fine mapping refines list of likely causal genes

We performed colocalization using the package ENLOC (19) to identify evidence of colocalization between GWAS and sQTL signals by calculating regional colocalization probabilities (RCP). Since ENLOC can only be applied to an intron region (LD block including the intron) in single tissues, for each gene, we calculated the maximum RCP (Max-RCP) across introns in the gene across the 11 tissues. We found single introns often had lower RCP compared to gene expression-based colocalization, therefore, we set a threshold of 0.10 for Max-RCPs. Genes with Max-RCP greater than the threshold are more likely to be causal. In our analysis, 88 of 249 genes had Max-RCP values greater than 0.1 (Supplementary Table 1). Overall, 56 genes exceeded both the fine-mapping and colocalization thresholds (Max-PIP greater than 0.80; Max-RCP greater than 0.10), exhibiting strong evidence of being causal genes (Supplementary Table 4, top 56 rows). Still, these genes need to be investigated in future functional experiments.

### Gene set enrichment and functional annotation corroborate splicing-TWAS results

Of the 249 genes identified by our joint splicing-TWASs, 216 are protein-coding genes, 19 are long non-coding RNA (lncRNA) genes, and 14 are pseudogenes. We tested the enrichment of the set of 235 protein coding and lncRNA genes against background gene sets from multiple databases using the FUMA software package (20). We limited the enrichment analysis to 33,527 background genes in FUMA. Three genes (*ENSG00000281357*, *ENSG00000284237*, *ENSG00000280670*) were not recognized in FUMA. We found the set of 235 genes identified by splicing-TWASs were significantly enriched in 42 background gene-sets at the threshold 0.05 for Bonferroni-adjusted p-values (Supplementary Table 5). These gene sets include a mammographic density set, an alcohol use disorder set, two body fat distribution sets (trunk fat ratio and leg fat ratio), and five breast related sets (breast cancer, estrogen-receptor negative breast cancer, breast size, NIKOLSKY BREAST CANCER 1Q21 AMPLICON, and NIKOLSKY BREAST CANCER 7Q21 Q22 AMPLICON). The enrichment in these gene sets suggests that the genes identified by splicing-TWASs may contribute to breast cancer etiology directly or through their impacts on known lifestyle/environmental risk factors. FUMA also identified differentially expressed gene (DEG) sets (genes which are significantly more or less expressed in a given tissue compared to others) for each of the 30 general tissues that FUMA selected from GTEx v8 data. The genes identified by splicing TWASs showed strong tissue specificity; for example, these genes are significantly enriched in the DEG sets in heart, pancreas, liver, blood, muscle, ovary, cervix uteri, and uterus tissues (Supplementary Figure 2).

## Discussion

In this study, we performed a multi-tissue and a breast-tissue-specific joint splicing-TWAS for overall breast cancer risk that combine information from multiple (excised) introns in a gene across multiple tissues (or in breast tissue only). We identified 249 significant genes. Among them, 88 genes in 62 loci have not been reported by previous TWASs; 17 genes in seven loci were at least 1 Mb away from previously published GWAS index variants and the remaining 232 genes are located known GWAS susceptibility loci. Of the 17 genes, 11 genes in 7 loci were not reported by previous TWASs.

As another focus of this study, we compared the results of two types of TWASs: splicing- and expression-TWASs. Our findings illustrated that multi-tissue and breast-tissue–specific joint splicing-TWASs identified genes that were not identified by the multi-tissue and breast-tissue– specific expression-TWAS when the two types of TWASs used the same summary statistics and prediction models trained in the same reference panel (GTEx v8). These findings suggested that sQTL-based splicing-TWASs may provide different information from eQTL-based expression-TWASs for breast cancer risk and may reveal new insights into genetic etiology of breast cancer.

We have checked in the literature the functional importance of the 11 genes (see Table 2) that are at least 1Mb away from published GWAS index SNPs and are not reported by previous TWASs. Here we briefly describe the importance of six genes, *TRIM4*, *GJC3*, *AZGP1*, *AFF1*, *SRP54*, and *ZNF* in cancer biology. Han et al (21) reported that *TRIM4* is downregulated in tamoxifen (TAM)LJresistant breast cancer cells, while the loss of TRIM4 is associated with an unfavorable prognosis; In vitro and in vivo experiments confirm that TRIM4 increased estrogen receptor alpha (ERLJ_α_) expression and the sensitivity of breast cancer cells to TAM.

*GJC3* and *AZGP1* are two genes also located at the same locus as *TRIM4* at 7q22.1, and the in-frame fusion of these genes (*AZGP1-GJC3*) has previously been reported in both triple-negative breast cancer and prostate cancer cells (22–24). This fusion event is a well-documented transcription-induced chimera (TIC). TICs occur when consecutive genes on a chromosome are spliced together, rendering their fusion product a functional protein. The intron-level significant association (P=8.23×10^-7^) in breast tissue of the same intron (intron_7_99968430_99971746) in both genes suggests that aberrant splicing in breast tissue could play a role in the development of this fusion in breast cancer.

*AFF1* is a proto-oncogene and member of the family of ALF transcription elongation factors located on chromosome 4 (25, 26). *AFF1* is translocated to chromosome 11 to fuse with *KMT2A* in nearly 50% of infant acute lymphoblastic leukemias (ALL). In these fusions, the transactivation domain of *AFF1* remains functional. Similarly, *AFF1*’s homolog, *AFF3* retains its transactivation domain when translocated in the minority of ALL *t(4;11)* translocations. Additionally, increased *AFF3* expression has been associated with tamoxifen resistance and breast cancer development in breast ductal acini cells(27, 28). More in-depth splicing quantification of RNA-seq in normal and malignant breast tissue is needed to elucidate the *AFF1* association with breast cancer.

*SRP* is a ribonucleoprotein with six subunits that targets proteins to the endoplasmic reticulum as they are translated (29), and in particular, *SRP54* has been shown to interact and decrease circulating copies of *TP53* in cervical cancer (30). *SRP54* has 23 documented splice variants. In our analysis, the splicing events intron_14_35001020_35007283 in this gene showed strong association with breast cancer in both the breast and adipose visceral omentum tissues (Table 2).

Another gene *ZNF18* at the 17p13.3 locus has been previously implicated in multiple cancer sites including breast cancer (31), diffuse large B cell lymphoma (32), clear cell endometrial carcinoma (33), and lung cancer(34). Interestingly, in lung cancer cell lines, overexpression of the tumor suppressor *MEN1* was shown to decrease the isoform abundance of *ZNF18* (34), suggesting that decreased expression of specific isoforms of *ZNF18* may play a role in carcinogenesis. Furthermore, the PIPs of intron excision events of *ZNF18* and *SRP54* in breast and several other tissues were high (PIP > 0.50, Supplementary Table 1) suggesting these genes may contain candidate causal isoforms that affect breast cancer risk.

Our splicing-TWASs can identify significant associations of non-causal introns and genes; this is similar to a GWAS, which can identify a susceptibility locus with a set of significant genetic variants, but cannot identify which variants in the locus are causal. The PIPs for individual introns in single tissues can provide useful information about how likely the corresponding genes are causal. However, compared to the PIP for a gene that was calculated based on an expression-TWAS, the PIPs for individual excised introns in a gene seemed relatively smaller on average. It is possible that a joint PIP combining information from multiple excised introns in a gene can be more useful for determining causal genes from the splicing-TWASs identified. We also noticed that the intron-based sQTL colocalization signal is weaker compared to the gene-based eQTL colocalization. We suggest a relatively small sQTL colocalization threshold 0.1 for the Max RCP to indicate association between corresponding gens and breast cancer.

We are not the first to attempt splicing-TWASs in the breast cancer context. He et al. (8) proposed an approach by integrating prior knowledge of susceptible transcription factor-occupied cis-regulatory elements (STFCREs) with TWAS (sTF-TWAS) in an effort to improve susceptible gene discovery. By applying their method to individual excised introns in the breast tissue in GTEx v8 and using the summary statistics of BCAC, He et al. performed a splicing-TWAS and identified 85 putative susceptibility genes for breast cancer at a threshold of 0.05 for Bonferroni adjusted p-values. In contrast, by using the same threshold for adjusted p-values, our multi-tissue splicing-TWAS and breast-tissue-specific joint splicing-TWAS identified 240 and 158 susceptible genes, respectively. Both of our multi-tissue and breast-tissue-specific joint splicing-TWAS analyses identified substantially more significant genes than He et al., possibly because of several notable differences in methodologies. First, we used GWAS data from a large number of breast cancer cases (N=133,511) and controls (N=291,090) combined from BCAC and UKB, while He et al. used the GWAS summary statistics of BCAC with a total of 122,977 cases and 105,974 controls. Second, for each gene, both our multi-tissue and breast-tissue– specific joint TWASs combined splicing-TWAS signals for multiple excised introns in the gene into one test, while He et al. performed multiple tests for the multiple excised introns, which increased the number of tests in the multiple testing correction and may have resulted in lower power. Third, our joint splicing-TWAS combined information across 11 tissues while He et al. only used the breast tissue from GTEx v8. Our results show that the multi-tissue approach identifies more genes compared to splicing-TWAS using breast tissue alone. This suggests that while breast tissue is an important tissue to utilize when conducting breast cancer splicing-TWASs, other tissues can contribute additional information for gene discovery. Fourth, we used splicing prediction models trained in GTEx v8 with the MASH method based on fine mapping to select possible causal sQTLs as predictors for each excised intron. Selecting possibly causal sQTL through fine mapping can reduce the probability that non-causal sQTLs were used in the prediction models (14). In addition, MASH can more accurately estimate the true sQTL effects (i.e., beta coefficients) on intron excision levels by jointly analyzing the sQTL summary statistics estimated in single tissues and accounting for correlation of non-zero sQTL effect sizes across the tissues; the estimates of beta coefficients of sQTLs by MASH were used as final weights in the splicing prediction models.

The current study has several limitations. First, although the multi-tissue joint splicing-TWAS identified more genes than breast-tissue-specific splicing-TWAS, it may have generated more false positive hits because: 1) it utilized other tissues that may not be truly causal to breast cancer (35), and 2) it used splicing prediction models trained in 11 tissues; the splicing prediction biases in any tissues may cause false positive findings. This last concern is mediated by the fact that the ACAT method used in our multi-tissue joint splicing-TWAS analysis calculates a weighted average of p-values from multiple tissues and is relatively conservative in identifying significant genes.

Second, the current study focused on overall breast cancer risk in women of European ancestry. We are currently working on splicing-TWASs that focus on ER-positive and ER-negative subtypes as well as intrinsic subtypes. In addition, future studies in other racial/ethnic populations are highly desirable. To date, RNA-seq data in the GTEx v8 have a small number of samples from non-European populations, creating a barrier to building accurate prediction models in these populations.

## Methods

### GWAS summary statistics and study population

We used results from a meta-analysis of GWAS summary statistics from BCAC GWAS and GWAS of breast cancer cases extracted from UK Biobank (UKB). BCAC GWAS is composed of 122,977 breast cancer cases and 105,974 controls. UKB GWAS includes 10,853 breast cancer cases and 262,614 controls. The details of UKB GWAS and meta-analysis are described in the methods of Gao et al.(5). We performed summary statistic imputation to optimize the accuracy of our GTEx splicing prediction models.

### Summary statistic-based imputation

For variants included in the GTEx prediction models but not in the GWAS summary statistics, we imputed z-scores with the method ImpG-Summary (36). The ImpG-Summary method estimates posterior mean of z-scores at unobserved SNPs based on the assumption that under the null hypothesis of no association, the vector ***Z*** of z-scores at all SNPs in a locus is approximately distributed as a Gaussian distribution, ***Z*** ∼ *N*(**0**, Σ), where Σ is the correlation matrix among all pairs of SNPs induced by LD. We used the GWAS summary statistics and correlation matrix estimated by using the genotype data in the GTEx samples as input of the ImpG-Summary method.

### Quantification of RNA splicing with LeafCutter

Li et al. (15) proposed an approach LeafCutter for the quantification of alternative splicing events by focusing on intron excisions (rather than whole isoform quantification). Leafcutter quantifies RNA splicing variation using short-read RNA-seq data. The core idea is to leverage spliced reads (reads that span an intron) to quantify (differential) intron usage across samples. Specifically, to identify alternatively excised introns, LeafCutter pools all mapped reads from a study and finds overlapping introns demarcated by split reads. LeafCutter then constructs a graph that connects all overlapping introns that share a donor or acceptor splice site. The connected components of the graph form clusters, which represent alternative intron excision events. Then LeafCutter estimate read proportions for all introns within alternatively excised intron clusters; the read proportions can be further standardized across individuals for each intron and quantile normalized across introns and then used as intron phenotype matrix for sQTL analysis or prediction model construction.

### Selection of tissues

For our multi-tissue joint splicing TWAS, we selected 11 tissues from the GTEx v8 data that are potentially relevant to breast cancer development or carcinogen metabolism (5), including female tissues (breast, ovary, uterus, and vagina), tissues that resemble connective and fat tissues in the breast (subcutaneous adipose, visceral adipose, and cultured fibroblasts), tissues related to immune cells (spleen, EBV-transformed lymphocytes, and whole blood), and liver.

### Intron splicing prediction models

Splicing prediction models were originally built in 49 tissues in GTEx (v8) samples of European ancestry that have the genotype and RNA-seq data; each of these 49 tissues has sample size in each tissue greater than 70. Sample size less than 70 may result in inaccuracy in prediction (10, 11, 14). Specifically, the prediction models were built with the following steps: 1) cis-sQTL analysis was performed by using fastQTL (37) in each tissue with the intron excision phenotypes (i.e., proportions standardized and then normalized by LeafCutter) (see previous section). For each intron excision event, all variants within the cis-window (±1Mb) with MAF>0.01 were considered and the following covariates were corrected in the linear regression models: sex, WGS platform, WGS library preparation protocol, top 5 genetic principal components, and PEER factors (10, 11). 2) Fine mapping was performed for each intron and its cis-region in each tissue by the dap-g method (19, 38) to select variants with minor allele frequency > 0.01 and posterior inclusion probabilities (PIPs) > 0.01 and to select excised introns with at least one credible set that had PIP > 0.1 (where the credible set PIP is sum of PIPs of variants in the set). Then in each credible set, only the variant with the highest PIP was kept. For the 49 tissues, a union of selected variants across 49 tissues was obtained and LD pruning was applied to the union of variants to remove redundant variants. 3) The multivariate adaptive shrinkage method (13) was used to estimate the true effects at the selected sQTL variants by jointly analyzing the marginal effect sizes and standard errors (SEs) of the sQTLs across the 49 tissues accounting for correlation among nonzero effects in different tissues (Barbeira, 2021 Genome Biology). 4) The predicted intron splicing level in each tissue was calculated as the linear combination of genotypes multiplying by their estimated effect sizes at the selected variants. In this study, we used the prediction models for 11 tissues potentially relevant to breast cancer. It is possible no prediction models could be constructed for some intron splicing events in some tissues because there are no strong sQTL signals for the intron phenotypes.

### Joint Splicing-TWAS test for multiple excised introns in a gene across multiple tissues

Suppose there are *J* excised introns in a gene with prediction models. The joint TWAS analysis generate a p-value for the gene by three steps: 1) performing traditional TWAS test for each intron in each of the 11 tissues by the software S-PrediXcan to obtain the p-values *p_jk_* (*j* = 1,…*J*; *k* = 1,…,11), where *j* denotes *j*-th intron and *k* denotes *k*-th tissue. 2) generating a tissue-specific p-value *p_ACAT,k_* for *k*-th tissue by constructing a tissue specific test statistic *T_ACAT,k_* with the ACAT method that combines p-values *p_jk_* (*j* = 1,…*J*) of all *J* introns with prediction models in the gene. Specifically, the ACAT test statistic is 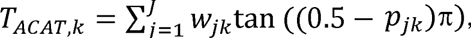, where w_jk_ are nonnegative weights. We used *w_jk_* = 1/*J*. The tissue specific p-value *p_ACAT,k_* of the ACAT test statistic is approximated by 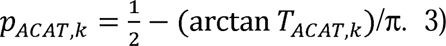 suppose the gene has p-values in *K* ≤11 tissues, we generate a joint p-value for the gene by the ACAT method again to combine the *K* tissue specific p-values *p_ACAT,k_* by a similar way as in step 2 except using weight =1/*K*.

### Conditional joint TWAS

To test if the signals at the 249 genes identified by our multi-tissue and breast-tissue-specific splicing-TWASs are independent of previously published GWAS index SNPs that were genome-wide significant (p<5×10^-8^), we performed splicing-TWASs that were conditional on these index SNPs. For each intron excision event, we defined two sets of SNPs: the target set of SNPs used for predicting the intron phenotype and the conditioning set of significant index SNPs from published GWASs within ±2 Mb of the transcription start or stop sites of the gene. By using the conditional and joint multiple-SNP (COJO) analysis method of Yang et al (39), for the target set of SNPs, we calculated adjusted effects (beta) on breast cancer risk and standard deviation of adjusted beta conditioning on the conditioning set of index SNPs. After performing COJO, we applied S-PrediXcan to these conditional summary statistics in single tissues and performed joint splicing TWAS to combine p-values from single introns in a gene and across individual tissues with the ACAT method.

### Intron-based colocalization analysis

For intron splicing events in the 249 genes that were identified by our splicing-TWASs, we calculated RCPs by the method ENLOC in each of the 11 tissues. ENLOC divides the genome into roughly independent LD blocks using the approach described in Berisa & Pickrell (40). For an intron located in a specific LD block, we calculated the colocalization probability of causal GWAS hits and causal sQTLs in the LD block by ENLOC. We used the GTEx (v8) sQTLs for the intron and the meta-analysis GWAS summary statistics in the LD block. For a gene with multiple introns, we assigned the maximum RCP across the 11 tissues as the gene-level RCP.

### Intron-based fine mapping

We performed intron-based statistical fine-mapping over the intron-trait association signals from S-PrediXcan using the software package FOCUS. For a LD block, we estimated a number of intron sets, each contained the causal introns at a predefined confidence level ρ (that is, ρ-credible gene sets; for example, ρ = 90%). We also computed the marginal PIP for each intron in the region to be causal given the observed TWAS statistics calculated from S-PrediXcan. FOCUS accounts for the correlation structure induced by LD and prediction weights used in the TWAS and controls for certain pleiotropic effects. FOCUS takes as input GWAS summary data, intron prediction weights, and LD among all SNPs in the LD region. We applied FOCUS to each of the 11 tissues and related splicing prediction weights from the GTEx v8. We assigned the maximum PIP of all introns across all tissues to a gene as gene-level PIP.

### Gene Set Enrichment and Functional Annotation

For the set of 249 significant genes identified by our splicing-TWASs, we conducted enrichment of 235 protein-coding and lncRNA genes against gene sets from multiple biological pathways, functional categories, and databases by the FUMA package. Specifically, we used the GENE2FUNC module of FUMA and specified 33,527 protein-coding and lncRNA genes as the background genes for enrichment testing. Multiple testing correction was performed per data source of tested gene sets (e.g., canonical pathways, GWAScatalog categories) using Bonferroni adjustment. We reported pathways/categories with adjusted p-value ≤ 0.05 and at least 2 genes that overlapped with the gene set of interest.

### Multi-tissue expression-TWAS

Our previous multi-tissue expression-TWAS (5) includes two steps: 1) performing a traditional TWAS analysis in each of the 11 tissues by the software S-PrediXcan to obtain the p-values *p_k_* (*k* = 1,…, 11), and 2) constructing test statistic by the ACAT method that combined p-values for each gene from the single tissue TWAS analyses across the 11 tissues. Gene expression prediction models were built with the genotype and RNA-seq data in 49 tissues of European ancestry from the GTEx project (v8) by a similar approach as described in the Section of Intron splicing prediction models and the prediction models for 11 tissues were used for the multi-tissue expression-TWASs (5). We used the summary statistics from meta-analysis of the BCAC GWAS and UKB GWAS results. Of the 19,274 genes tested in our joint expression-TWAS analysis, we identified 299 genes whose predicted expression was associated with breast cancer risk at the Bonferroni-corrected significance level (p < 2.59×10^-6^). Only 141 genes were identified when TWAS analysis used only breast tissue, i.e. conventional single-tissue TWAS approach. Of these 141 genes, 131 genes were also identified in the multi-tissue TWAS. The remaining 10 genes identified only in the breast-tissue TWAS analysis were also marginally significant in the multi-tissue TWAS (p < 0.05), so we considered the 309 genes from either expression-TWASs in this study for comparison with the results of splicing-TWASs.

## Figure and Table Legends

**Supplementary** Figure 1. Comparison of joint splicing-TWAS and joint expression-TWAS Manhattan plots.

**Supplementary** Figure 2. Differential analysis of expression of the splicing-TWAS identified genes in GTEx v8 shows tissue specificity. Significantly enriched differentially expressed gene sets (Bonferoni adjusted p < 0.05) are highlighted in red. The P values were from hypergeometric test.

**Supplementary Table 1.** The 249 genes identified by multi-tissue or breast-tissue-specific joint splicing-TWAS.

**Supplementary Table 2.** The 158 genes identified by breast-tissue-specific splicing-TWAS.

**Supplementary Table 3.** The 110 genes identified by our multi-tissue or breast-tissue-specific splicing-TWAS but not by our previous multi-tissue or breast-tissue-specific expression-TWAS using the same GWAS summary statistics.

**Supplementary Table 4.** The 114 candidate causal genes identified by fine-mapping analysis (sorted by Max RCP).

**Supplementary Table 5.** Significant gene sets in the enrichment analysis using FUMA.

## Data Availability

In this study, we only used existing datasets that are publicly available (see the section of Web resources). The code pipeline and results for our multi-tissue joint splicing-TWAS analysis will be available at https://zenodo.org/. For specific method code, we made minor modifications to S-PrediXcan to combine results with ACAT (https://github.com/shugamoe/MetaXcan/tree/catch_up). We also made minor modifications to FOCUS to accommodate PrediXcan GTEx v8 MASHR models (https://github.com/shugamoe/focus).

## Acknowledgements

This work was supported by the National Cancer Institute (R01 CA242929, R01 CA228198, P20 CA233307), Breast Cancer Research Foundation (BCRF-22-071), National Institutes of Health (R01 MD013452), and the NIDDK (P30 DK20595). For BCAC data, the breast cancer genome-wide association analyses were supported by the Government of Canada through Genome Canada and the Canadian Institutes of Health Research, the ‘Ministère de l’Économie, de la Science et de l’Innovation du Québec’ through Genome Québec and grant PSR-SIIRI-701, The National Institutes of Health (U19 CA148065, X01HG007492), Cancer Research UK (C1287/A10118, C1287/A16563, C1287/A10710) and The European Union (HEALTH-F2-2009-223175 and H2020 633784 and 634935). All studies and funders are listed in Michailidou et al (Nature, 2017). We thank Yang Li for consulting on quantification of intron excisions and Sarah Sumner for help editing the paper.

## Author contributions

G.G., D.H., H.K.I. conceived the study and contributed to the study design. J.M., G.G., A.N.B., P.N.F., Z.M., D.H., and H.K.I. performed data analyses. G.G., and P.N.F. wrote the first version of the manuscript. G.G., J.M., A.N.B., P.N.F., J.L.L., O.I.O., D.H., and H.K.I revised the manuscript.

## Web resources

PrediXcan GTEx v8 MASHR models, https://predictdb.org/; Summary statistics of meta-analysis of BCAC and UKB, https://zenodo.org/record/7814694#.ZDaspXbMK5d; FUMA software, http://fuma.ctglab.nl; COJO (GCTA), https://yanglab.westlake.edu.cn/software/gcta/; Enloc, https://github.com/xqwen/integrative; S-PrediXcan, https://github.com/hakyimlab/MetaXcan and https://github.com/hakyimlab/summary-gwas-imputation; UK Biobank, http://ukbiobank.ac.uk

