## Supplemental figures 1-2 for "A splicing-based multi-tissue joint transcriptome-wide association study identifies susceptibility genes for breast cancer"

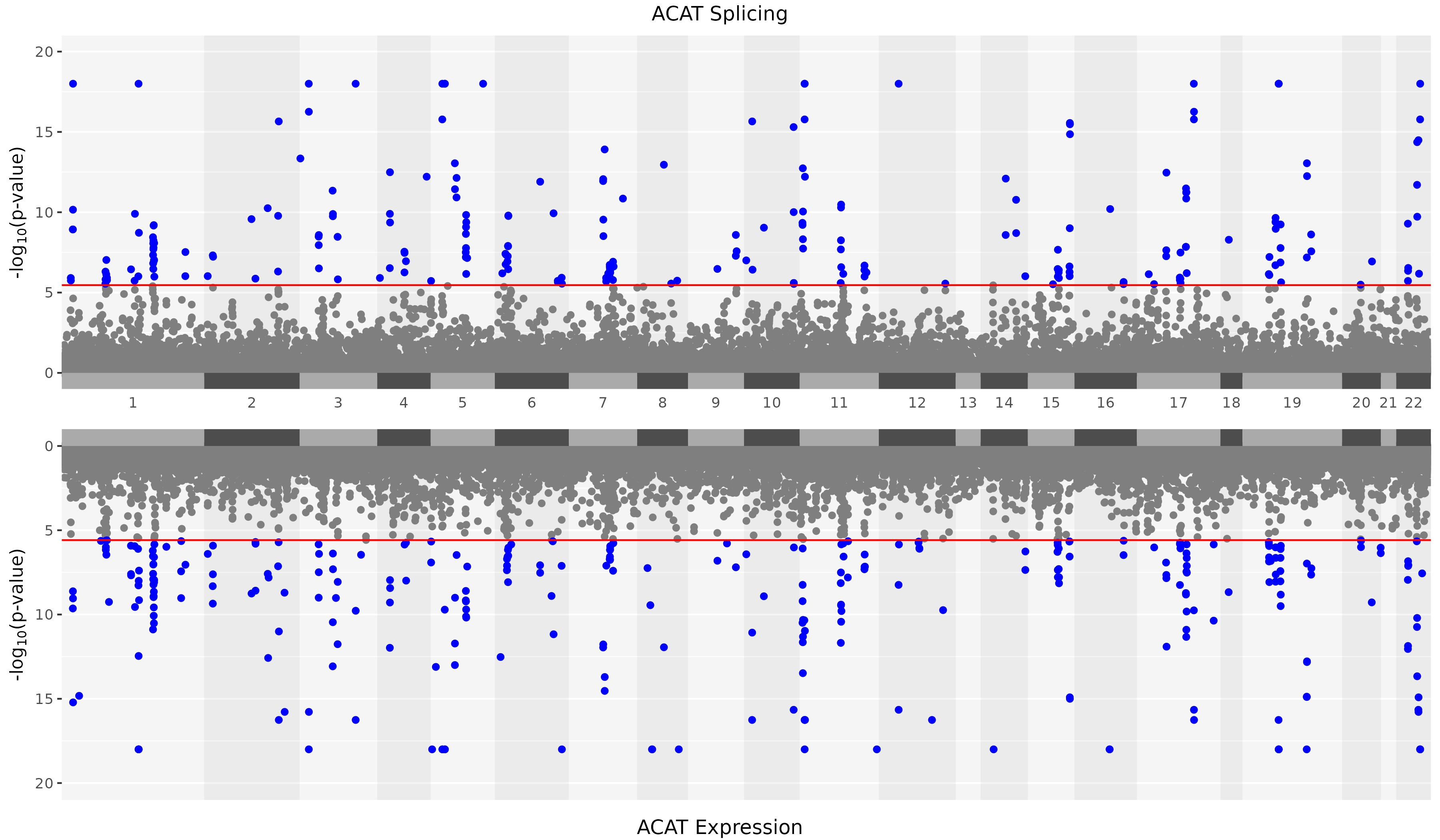
Supplementary Figure 1. Comparison of multi-tissue joint splicing-TWAS and multi-tissue expression-TWAS Manhattan plots. The top figure is the multi-tissue splicing-TWAS Manhattan plot. Each point corresponds to an ACAT association test combining S-PrediXcan p-values of introns in a gene across 11 tissues. The red line represents the boundary for splicing-TWAS significance level (4.58 × 10^−7^). The bottom figure is the multi-tissue expression-TWAS Manhattan plot where each point is the result of an ACAT association test combining S-PrediXcan p-values at a gene across 11 tissues. The red line corresponds to transcriptome-wide significance for expression (4.58 × 10^−7^).

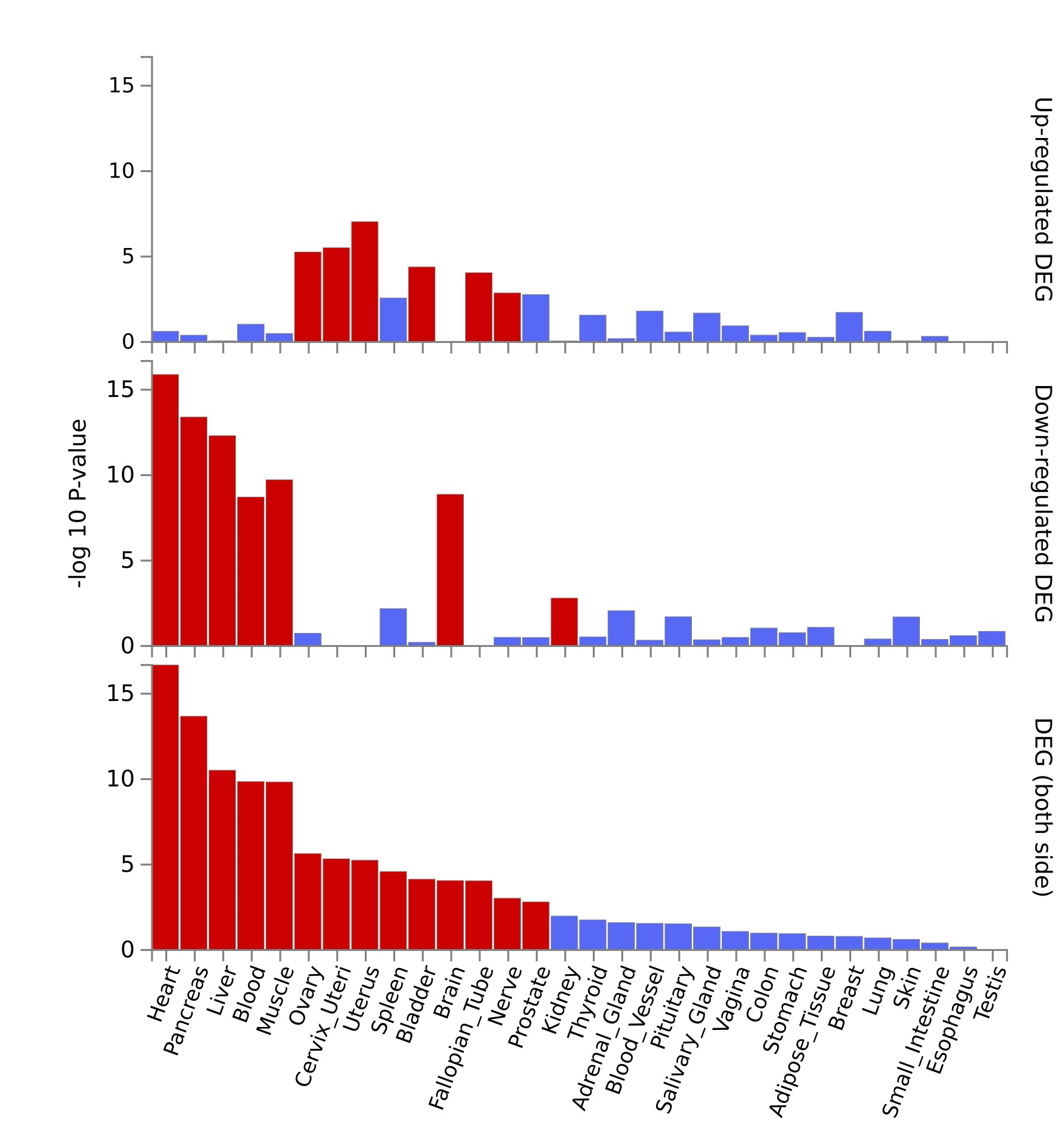

**Supplementary Figure 2.** Differential analysis of expression of the splicing-TWAS identified genes in GTEx v8 shows tissue specificity. Significantly enriched differentially expressed gene sets (Bonferoni adjusted p < 0.05) are highlighted in red. The P values were from hypergeometric test.
